# Assessment of a Diagnostic Strategy Based on Chest Computed Tomography in Patients Hospitalized for COVID-19 Pneumonia: an observational study

**DOI:** 10.1101/2020.06.29.20140129

**Authors:** Marine Thieux, Anne-Charlotte Kalenderian, Aurélie Chabrol, Laurent Gendt, Emma Giraudier, Hervé Lelievre, Samir Lounis, Yves Mataix, Emeline Moderni, Laetitia Paradisi, Guillaume Ranchon, Carlos El Khoury, on behalf of MHM Research Group

## Abstract

**Objectives:** To assess the relevance of a diagnostic strategy for COVID-19 based on chest computed tomography (CT) in patients with hospitalization criteria.

**Setting:** Observational study with retrospective analysis in a French emergency department (ED).

**Participants and intervention:** From March 3 to April 2, 2020, 385 adult patients presenting to the ED for suspected COVID-19 underwent an evaluation that included history, physical examination, blood tests, real-time reverse transcription-polymerase chain reaction (RT-PCR) and chest CT. When the time-interval between chest CT and RT-PCR assays was longer than 7 days, patients were excluded from the study. Only patients with hospitalization criteria were included. Diagnosis accuracy was assessed using the sensitivity and specificity of RT-PCR.

**Outcomes:** Sensitivity and specificity of RT-PCR, chest CT (also accompanied by lymphopenia) were measured and were also analyzed by subgroups of age and sex.

**Results:** Among 377 included subjects, RT-PCR was positive in 36%, while chest CT was compatible with a COVID-19 diagnosis in 59%. In the population with positive RT-PCR, there were more men (55% vs 37%, p=0.015), a higher frequency of reticular and, or, interlobular septal thickening (53% vs 31%, p=0.02) as well as a higher frequency of bilateral lesion distribution (98% vs 86%, p=0.01) compared to the population with negative RT-PCR. The proportion of lymphopenia was higher in men vs women (47% vs 39%, p=0.03) and varies between patients >80 versus 50-80 and p<0.001).

Using CT as reference, RT-PCR obtained a sensitivity of 61%, specificity of 93%. There was a significant difference between CT and RT-PCR diagnosis performance (p*<*0.001). When CT was accompanied by lymphopenia, sensitivity and specificity of RT-PCR were respectively 71% and 94%. CT abnormalities and lymphopenia provided diagnosis in 29% of patients with negative PCR.

**Conclusions:** Chest CT had a superior yield than RT-PCR in COVID-19 hospitalized patients, especially when accompanied by lymphopenia.

## INTRODUCTION

In December 2019, a new coronavirus was identified after several cases of pneumonia of “unknown cause” [1] emerged in Wuhan, China. This novel coronavirus, named SARS-CoV-2 (Severe Acute Respiratory Syndrome Coronavirus 2),[2] is responsible for COVID-19 infection. The World Health Organization (WHO) declared the COVID-19 outbreak a global pandemic on March 11, 2020.[3] As of May 11, 2020, first day post-lockdown, more than 4.2 million confirmed cases of COVID-19 have been reported worldwide, with more than 291 500 deaths. Our country is the sixth most affected European country by COVID-19 with over 140 100 cases and slightly less than 27 000 deaths.[4]

In this context, an unusually high volume of patients presenting to emergency departments was expected. To face this situation, there was a need for emergency physicians to act quickly and accurately in the care of COVID-19 patients, which necessitates to anticipate the use of hospital resources. The challenge was to ensure fluid management by implementing a reliable screening of patients, in order to distinguish those infected with the virus from non-infected ones, and to place them in the right pathway of care without delay. In the case of the COVID-19 outbreak, biologists and radiologists played an important role in the triage and the diagnostic strategy.[5]

While a real-time reverse transcription-polymerase chain reaction (RT-PCR), consisting of a throat swab sample, has been described as the gold standard method for COVID-19 diagnosis,[5] its limited sensitivity and long time-to-result [6-8] prompted us to look for a faster and more effective diagnostic test.

Faced with these considerations, our emergency department decided early on to implement a diagnostic strategy based on chest CT for suspected COVID-19 patients with hospitalization criteria. The objective was to identify radiographic features consistent with COVID-19 pneumonia in patients considered with a high clinical probability of COVID-19 and to assess the severity of lung lesions without delay. Knowing the low specificity of CT in this indication,[7-9] it was associated with blood tests. This approach was intended to improve the speed of the diagnostic decision-making, hence allowing a faster identification of infected patients.

The purpose of this study was to evaluate the relevance of a COVID-19 diagnostic strategy based on chest CT findings in patients with hospitalization criteria.

## METHODS

This study was approved by a local Ethics Committee and the data treatment was approved by the National Commission for Liberties and Data Protection (CNIL) (number 2217103 v 0).

### Patients

Each patient suspected of novel coronavirus infection went over an evaluation based on a written protocol that included medical history and vital-parameter measurement by two dedicated nurses in the emergency department. Patients with hospitalization criteria (i.e., respiratory rate >20/min, pulse oximetry [SpO2] <93%, systolic blood pressure [SBP] <100 mmHg) underwent chest CT, RT-PCR assay and blood tests, and were directly admitted to a dedicated hospitalization unit. Before transferring a patient to a dedicated COVID-19 medical unit, the diagnosis of COVID-19 pneumonia was confirmed by agreement between emergency physicians, pneumologists and radiologists, according to the test results.

From March 3 to April 2, 2020, 385 consecutive adult patients with hospitalization criteria who underwent both chest CT and RT-PCR were retrospectively enrolled in our non-academic emergency department receiving more than 50.000 patients per year. For each patient, age, sex, RT-PCR and chest CT results, the delay between these examinations (in days) and laboratory results were collected from medical records. Patients with no criteria for hospitalization did not have CT nor blood tests and were not included in this study.

### RT-PCR

The RT-PCR assays were performed by sets, three times a day (8 am, 1 pm, 5 pm), using a multiplexed real-time PCR (EBX 041 SARS CoV2 kit by Eurobio Scientific). This kit follows the design recommended by the French National Reference Center (CNR) and allows the detection of the 3 identification genes of the SARS-CoV-2 virus (i.e., N gene, RdRp gene, E gene) as recommended by the WHO. [10]

### Chest CT protocols

Immediately after the RT-PCR swabs, patients underwent chest CT to determine the presence or absence of viral pneumonia. All chest CT acquisitions were obtained with the patients in supine position during end-inspiration without contrast medium injection. Chest CT performed on a 128-slice CT (GE Revolution EVO 128 Slice CT Scanner, GE Medical Systems, Milwaukee, WI, USA) dedicated only to patients with suspected COVID-19. The following technical parameters were used: tube voltage: 100 kV; tube current modulation 80-120 mAs; spiral pitch factor: 0.9131; collimation width: 0.6.

### Image analysis

All CT images were independently reviewed by two radiologists (AC and ACK with 13 and 15 years of experience in interpreting CT images, respectively), blinded to the RT-PCR results and to the first interpretation. They both described main CT features then classified chest CTs according to their compatibility with COVID-19 diagnosis (**Figure 1**). After evaluation, any disagreements were resolved by discussion and consensus.

**Figure 1.**
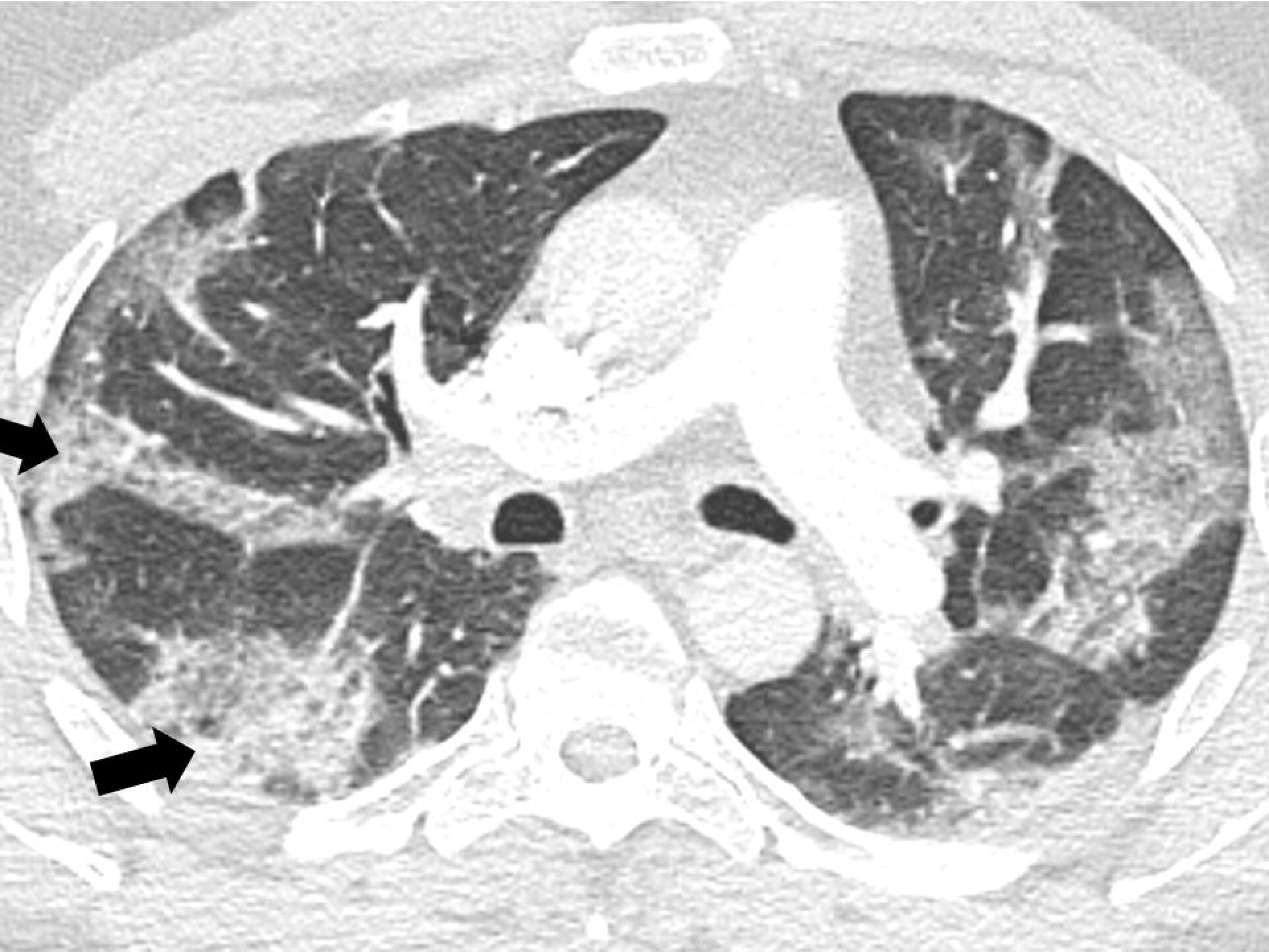
Chest CT showing consolidation with thickened intralobular and interlobular septum (black arrow) segmental and subpleural consolidation in both lungs.

For patients with multiple RT-PCR assays or multiple chest CT, only the closest evaluation between chest CT and RT-PCR assay, in terms of time interval, was analyzed.

### Biological data

The blood samples, including C-Reactive Protein (CRP) and lymphocyte value, were collected the day of the hospitalization. Formula count and CRP were among those routinely taken. Normal lymphocyte value was ranging from 1 to 3.9 G/L (lymphopenia in adults was defined as count of <1 G/L). Cutoff value for CRP was >5 mg/L. Results were then classified as pathologic or not according to these reference values. For patients with multiple samples, the most pathological value between the first and second day of hospitalization was selected.

### Statistical analysis

The statistical analysis was conducted by MT using R (R CORE Team). [11] Continuous measures were expressed as medians and interquartile ranges (IQR), while categorical measures were reported as counts and percentages. Using CT results as reference associated or not with laboratory measures, the sensitivity, specificity, positive predictive value (PPV) and negative predictive value (NPV) of RT-PCR were calculated and compared using Chi2 test. These values were also analyzed by subgroups of age (i.e., <50 years old, between 50 and 80 years old and >80 years old) and sex to evaluate the performance of diagnostic tests in these subgroups. Recent studies indicate that patients ≥50 years old are at higher risk than the youngest, [12] and the mortality rate of patients >80 years in critical care units is close to 100%.[13] Lastly, imaging and laboratory abnormalities may vary according to age.[7,14,15]

Comparisons between groups were realized using Wilcoxon, Kruskal-Wallis, Fisher’s exact and Chi2 tests when adapted. Statistical analyses were conducted with a 95% confidence interval. Missing values were removed from the analyses.

## RESULTS

### Patients results

From March 3 to April 2, 2020, 385 consecutive adult patients with hospitalization criteria underwent both RT-PCR and CT. Among them, 377 (98%) patients had both tests less than 7 days apart. Patients were excluded from the analysis when the time-interval between chest CT and RT-PCR assay was longer than 7 days (n=8) as suggested by Ai et al.[6] Their characteristics are summarized in **Table 1**. Forty-six percent were men (median age: 73,1 years old) and 54% were women (median age: 70,1 years old). All of them were hospitalized in a dedicated unit. RT-PCR was positive in 136 (36%) of them whereas CT was consistent with COVID-19 in 223 (59%). Among the latter, 126 (57%) were associated to a positive RT-PCR and 18 (8%) had “inconclusive” RT-PCR results.

**Table 1.** Clinical characteristics, diagnostic tests and laboratory findings of patients with coronavirus disease 2019 (COVID-19) pneumonia.

| **Clinical characteristics** | **All patients (n = 377)** |
| --- | --- |
| Men (%) | 173 (46%) |
| Age, median [IQR] | 72.1 [28.5] |
| Hospitalization (%) | 377 (100%) |
| **Diagnostic tests** |  |
| RT-PCR positive (%) | 136 (36%) |
| CT compatible (%) | 223 (59%) |
| PCR - CT delay, days, median [IQR] | 0 [0] |
| **Laboratory results** |  |
| Lymphopenia (%) | 152 (45%) (NA = 37) |
| Lymphocytes, median [IQR] | 1.06 [0.8] (NA = 37) |
| Elevated CRP (%) | 285 (86%) (NA = 44) |
| CRP, median [IQR] | 69.1 [140.8] (NA = 44) |

IQR, Interquartile Range; NA, not available.

Sex comparison indicates that lymphopenia and elevated CRP levels were more frequently observed among men (47% vs 39% [p*=*0.03] and, 91% vs 81% [p*=*0.008], respectively). There were no statistically significant differences between men and women in age, RT-PCR or CT results.

Among patients with COVID-19 pneumonia, 20.4% were <50 years old, 46.4% were 50-80 years old and 33.2% were >80 years old. Comparison between age groups suggested differences in the rate of positive RT-PCR and CT (p<0.001 and p=0.042, respectively) (**Table 2**). Two-by-two comparison analyses showed that RT-PCR was more frequently positive in the 50-80 year-old-group compared to the group <50 years old (44% vs 22%, p<0.001) but no significant difference was found in comparison to the group >80 years old (44% vs 34%, p*=*0.16).

**Table 2.** Comparison between clinical characteristics, diagnostic tests and laboratory findings according to <50, 50-80 and >80 years old patients with coronavirus disease 2019 (COVID-19) pneumonia.

|  | <50 years (n = 77) | 50 - 80 years (n = 175) | > 80 years (n = 125) | *P* |
| --- | --- | --- | --- | --- |
| **Clinical characteristics** |  |  |  |  |
| Men, (%) | 30 (39%) | 83 (47%) | 60 (48%) | 0.391 |
| Age, median [IQR] | 39 [9.5] | 68.3 [14.6] | 86.6 [7] | **<0.001** |
| Hospitalization (%) | 77 (100%) | 175 (100%) | 125 (100%) | 1.00 |
| **Diagnostic tests** |  |  |  |  |
| PCR positive (%) | 17 (22%) | 77 (44%) | 42 (34%) | **<0.001** |
| CT compatible (%) | 36 (47%) | 111 (63%) | 76 (61%) | **0.042** |
| PCR - CT delay, days, median [IQR] | 0 [1] | 0 [0] | 0 [0] | 0.438 |
| **Laboratory results** |  |  |  |  |
| Lymphopenia (%) | 13 (22%) (NA = 18) | 68 (43%) (NA = 15) | 71 (59%) (NA = 4) | **<0.001** |
| Lymphocytes, median [IQR] | 1.36 [1.05] (NA = 18) | 1.16 [0.74] (NA = 15) | 0.87 [0.59] (NA = 4) | 0.223 |
| Elevated CRP (%) | 38 (67%) (NA = 20) | 138 (79%) (NA = 17) | 109 (92%) (NA = 7) | **< 0.001** |
| CRP, median [IQR] | 26 [116.9] (NA = 20) | 71.5 [135.5] (NA = 17) | 87.6 [133.54] (NA = 7) | 0.420 |

IQR; Interquartile Range, NA; not available.

Moreover, the group >80 years old also had more frequent positive RT-PCR than the group <50 years old (34% vs 22%, p<0.001). Additionally, two-by-two comparisons showed that CT was more frequently positive in the group of patients 50-80 years old than in the group <50 years old (63% vs 47%, p*=*0.02), without significant difference with the group >80 years old (63% vs 61%, p*=*0.72). The group of patients >80 years old is more likely to have a positive CT than the group <50 years (61% vs 47%, p=0.059) but this result was not statistically significant.

The three-group comparison also showed that lymphopenia tended to be more present in patients >80 years old than those 50-80 years old (59% vs 43%, p*=*0.01) or <50 years old (59% vs 22%, p<0.001). Patients 50-80 years old were also more likely to have lymphopenia than those <50 years old (43% vs 22%, p*=*0.01). In addition, patients >80 years old tended to have higher levels of CRP than patients <50 years old (92% vs 67%, p<0.001) but they were not significantly different from patients 50-80 years old (92% vs 79%, p*=*0.234). Lastly, patients 50-80 years old were more likely to have elevated CRP levels than those <50 years old (79% vs 67%, p=0.001).

### Diagnostic tests results

All 223 patients (except one) with COVID-19 CT compatible had ground-glass opacities (GGOs) and 64.1% had consolidation. Bilateral distribution of CT abnormalities was observed in 94.2% of cases (**Table 3**).

**Table 3.** Imaging features in patients with a positive CT (n=223)

| Characteristics | COVID-19 CT compatible |
| --- | --- |
| Men (%) | 105 (45%) |
| Age, median [IQR] | 72.2 [26] |
| Ground-glass opacity (%) | 222 (99.6%) |
| Consolidation (%) | 143 (64.1%) |
| Reticulation – Thickened interlobular septa (%) | 100 (45.9%) (NA = 5) |
| Nodules (%) | 15 (6.7%) |
| Lesion distribution (%) |  |
| *Left* | 4 (1.8%) |
| *Right* | 9 (4%) |
| *Bilateral* | 210 (94.2%) |
| Impairment extent, median [IQR] | 25 [35] |
| Pleural effusion (%) | 34 (18.5%) (NA = 39) |
| Mucocele (%) | 9 (4.9%) (NA = 39) |

IQR; Interquartile Range, NA; not available.

Comparing patients with positive RT-PCR and negative RT-PCR patients (**Table 4**), we found that the former group included more men (p*=*0.015) and presented more often reticulation and, or, thickened interlobular, bilateral distribution of imaging abnormalities and more extensive lesions at CT.

**Table 4.**
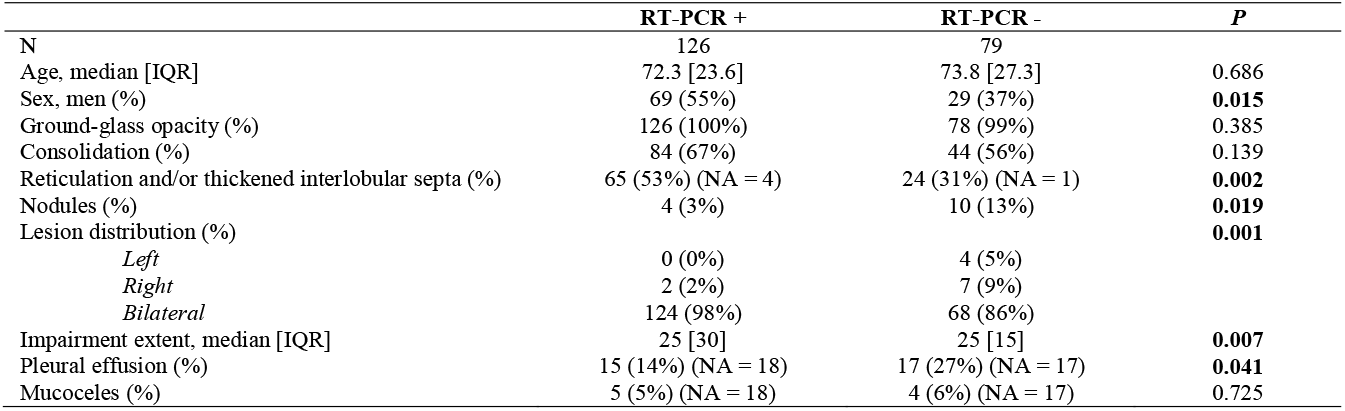
Positive CT characteristics (n=223; 18 inconclusive polymerase chain reaction [PCR]) comparison between PCR + vs PCR -

|  | RT-PCR + | RT-PCR - | *P* |
| --- | --- | --- | --- |
| N | 126 | 79 |  |
| Age, median [IQR] | 72.3 [23.6] | 73.8 [27.3] | 0.686 |
| Sex, men (%) | 69 (55%) | 29 (37%) | **0.015** |
| Ground-glass opacity (%) | 126 (100%) | 78 (99%) | 0.385 |
| Consolidation (%) | 84 (67%) | 44 (56%) | 0.139 |
| Reticulation and/or thickened interlobular septa (%) | 65 (53%) (NA = 4) | 24 (31%) (NA = 1) | **0.002** |
| Nodules (%) | 4 (3%) | 10 (13%) | **0.019** |
| Lesion distribution (%) |  |  | **0.001** |
| *Left* | 0 (0%) | 4 (5%) |  |
| *Right* | 2 (2%) | 7 (9%) |  |
| *Bilateral* | 124 (98%) | 68 (86%) |  |
| Impairment extent, median [IQR] | 25 [30] | 25 [15] | **0.007** |
| Pleural effusion (%) | 15 (14%) (NA = 18) | 17 (27%) (NA = 17) | **0.041** |
| Mucocele (%) | 5 (5%) (NA = 18) | 4 (6%) (NA = 17) | 0.725 |

There was no significant difference in the rate of lymphopenia between the group with a CT compatible compared to the group with a positive RT-PCR (52% vs 59%, p*=*0.217).

### Diagnostic accuracy

All diagnostic tests results are illustrated **Figure 2**.

**Figure 2.**
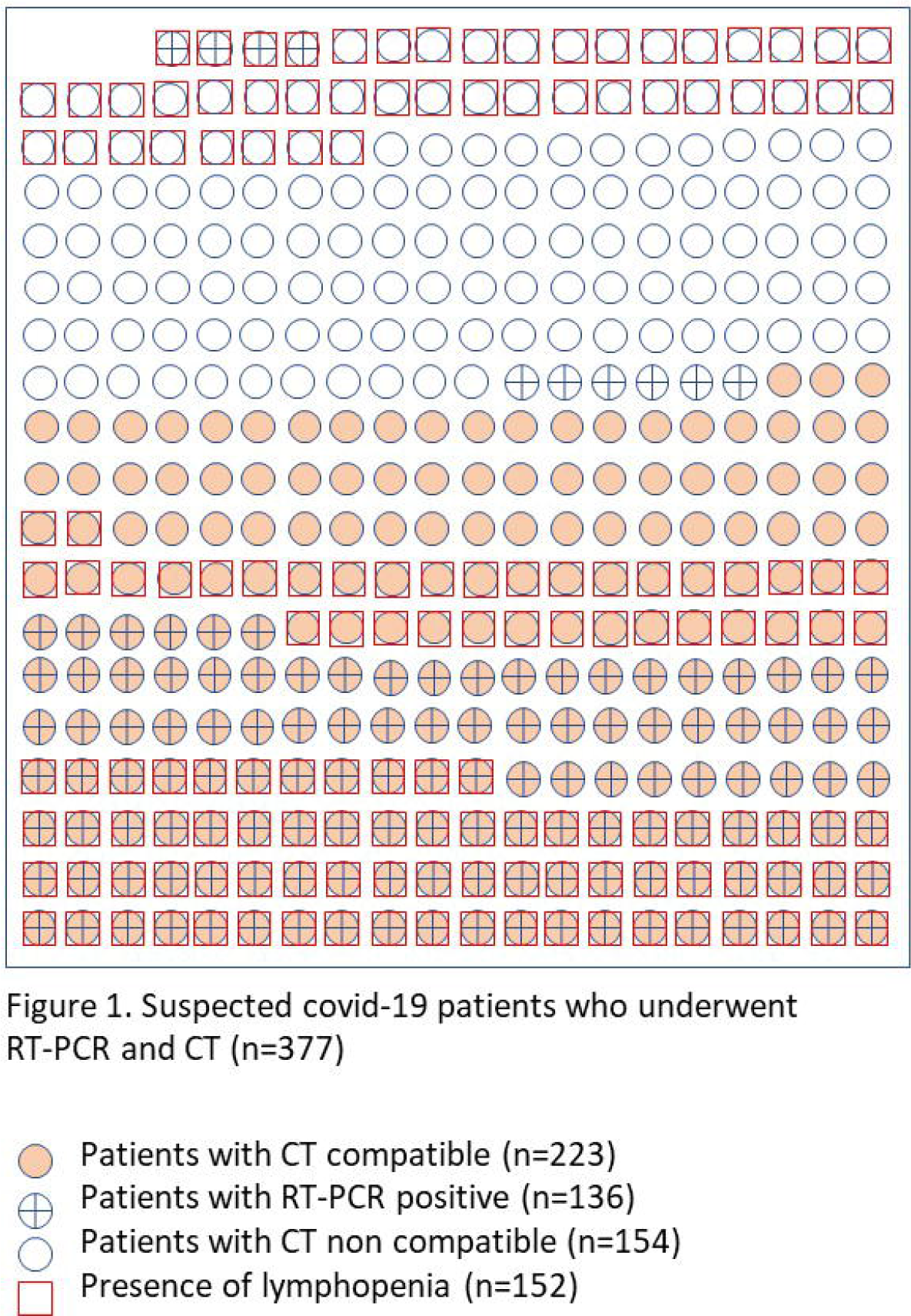
Adapted from Tze-Wey Loong (2003).[16]

When using CT as reference, values of sensitivity of 61%, specificity of 93%, PPV of 93% and NPV of 62% were obtained for RT-PCR. There was a significant difference between CT and RT-PCR diagnosis performance (p<0.001).

When using CT associated with lymphopenia as reference, values of sensitivity of 71%, specificity of 94%, PPV of 93% and NPV of 73% were obtained for RT-PCR. There was a significant difference between CT accompanied by lymphopenia and RT-PCR diagnostic performance (p<0.001).

In men, when using CT as reference, values of sensitivity of 70%, specificity of 97%, PPV of 97% and NPV of 67% were obtained for RT-PCR. There was a significant difference between CT and RT-PCR diagnostic performance in men (p<0.001). In women, values of sensitivity and specificity for RT-PCR decreased, 53% and 90%, respectively, PPV 80% and NPV of 58%, respectively. There was a significant difference between CT and RT-PCR diagnostic performance in women (p<0.001).

In groups of age, values of RT-PCR sensitivity for patients <50, 50-80 and >80 years old were 54%, 69% and 54%, respectively. Values for specificity were 94%, 92% and 94%, respectively. Values for PPV were 88%, 94%, and 93%, respectively and values for NPV were 69%, 63% and 58%, respectively.

There was a significant difference between CT and RT-PCR diagnostic performance in the <50 year-old-group (p*=*0.001), as well as in the 50 to 80 year-old-group (p<0.001) and in the >80 year-old-group (p<0.001).

## DISCUSSION

In the context of coronavirus disease 2019 pandemic, early diagnosis was crucial for disease treatment and control in order to avoid the overcrowding of the emergency departments and the spreading of the virus.[9]**Error! Bookmark not defined**.

While the real-time reverse-transcription polymerase chain reaction is considered as the gold standard for the diagnosis of COVID-19, its long time-to-results,[5] and lack of sensitivity,[6,17,18] prevent emergency physicians to be efficient in the initial patient triage. In a recent study, chest CT was proved to be more efficient in the diagnosis of COVID-19 than RT-PCR, with a greater sensitivity and rapidity. Moreover, it was reported to be a very useful tool to detect COVID-19 features and to assess the severity of lung lesions.[5,6,9,19,20]

On this basis, emergency physicians have agreed with radiologists and biologists on a local algorithm, based on using both RT-PCR and chest CT along with blood tests for an efficient diagnostic strategy.

In order to optimize the sensitivity of CT, we chose to perform this test only on patients with hospitalization criteria. We also searched for lymphopenia to improve its specificity.

In this study, 223/377 (59%) patients had evidence of CT abnormalities compatible with COVID-19 pneumonia and 136/377 (36%) a positive RT-PCR. With CT results as reference, our analysis shows that the sensitivity and specificity of RT-PCR to detect COVID-19 were 61% and 93% respectively when both tests were performed on all patients the same day, which is consistent with the results of recent studies.[6,18]

In clinical practice, the purpose of all diagnostic tests is to reduce clinical uncertainty. However, the low sensitivity rate of RT-PCR could lead to false negatives,[21] thus indicating that negative RT-PCR tests must be cautiously interpreted. This uneven sensitivity may be caused by immature development of RT-PCR technology, low viral load in the patient or improper clinical sampling.[17-19]

Based on data published in recent literature, the typical CT features were similar in almost all patients.[6] In this study, the clinical history, the CT features and the lesion distribution were determining factors for radiologists when interpreting whether a CT was consistent with COVID-19 or not. GGOs were the most common abnormality (99.6%), followed by consolidation (64.1%) and reticulation and, or, thickened interlobular septa (45.9%). Lastly, the lesion distribution was predominantly bilateral, in 94.2% cases.

Interestingly, when comparing the results according to sex and age of patients, we noted that men tended to be more affected than women.[1,22] Additionally, older patients were more often diagnosed with GGOs than younger patients.[14] This finding may be due to the median age of the patients (72.1 years) enrolled in the study, as GGOs are also naturally present in the elderly. Moreover, this may also be linked to the inclusion criteria of this study as GGOs are most typically found in patients with mild to severe COVID-19 pneumonia.[23] Furthermore, in a recent study, it has been suggested that the size and type of CT abnormalities were related to the severity of the disease.[24]

However, as COVID-19 pneumonia shares CT features with other conditions, such as influenza,[7,8,9] CT is thereby unlikely a reliable standalone tool to confirm or rule out COVID-19 infection.[25]

To address these issues, CT was combined with blood tests in addition to RT-PCR to improve the accuracy of the diagnosis. Several studies reported that elevated CRP levels and decreased lymphocyte counts were commonly found in patients with typical symptoms of COVID-19 and radiological abnormalities.[23,26,27] Other studies found that the proportions of normal or decreased leukocyte counts and decreased lymphocytes in the COVID-19 group were higher than those of the control group.[21]

In our study, values of sensitivity and specificity for RT-PCR were higher when CT was associated with lymphopenia (respectively 71% and 94%, versus 61% and 93%). In addition, 30% of patients with chest CT consistent with COVID-19 infection but negative RT-PCR were associated with lymphopenia.

Subgroup analysis emphasizes lower sensitivity of RT-PCR in patients <50 years old and those >80 years old, as well as in women.

While our study provided an overview of real-life professional practices, time and resources management during a health crisis, it has some limitations. First, the prevalence of the disease is unknown. In absence of mass screening, we believe that it is much higher than announced, which may change positive and negative predictive values and impacts the clinical decision.[16]

Secondly, sensitivity and specificity values can be subject to bias, which we aimed to reduce by selecting patients with hospitalization criteria. On the other hand, we are aware that our population was not representative of all COVID-19 patients.

Finally, because our study was observational, some data are missing.

## CONCLUSION

This study showed that CT sensitivity is superior to that of reverse-transcription polymerase chain reaction in patients with hospitalization criteria, especially when accompanied by lymphopenia. Its availability and rapidity to deliver result make it the most appropriate test for the emergency departments.

## Data Availability

All data relevant to the study is not in a repository (deidentified participant data).

## Acknowledgments

Authors are thankful to all physicians and nurses who have contributed to include patients in the local protocol.

## Contributors

This study was conceived by Marine THIEUX, MSc, Anne-Charlotte KALENDERIAN, MD, Aurélie CHABROL, MD, Laurent GENDT, Hervé LELIEVRE, Samir LOUNIS, Emma GIRAUDIER, MSc, Yves MATAIX, MD, Emeline MODERNI, MSc, Laetitia PARADISI, PharmD, Guillaume RANCHON, MD, & Carlos El KHOURY, MD, PhD. All the authors participated to the conception, design, writing and/or contributed to the critical process of the manuscript. MT carried out the statistical analysis and drafted the article, CEk, EG and EM drafted the article, AC and ACK interpreted CT, CEk conceived and designed the study, AC, LG, HL, SL, YM, LP, ACK and GR validated the clinical relevance, read and brought critical content to the study. All of authors approved the current version of the paper.

## Patient consent

Not required

## Data sharing statement

All data relevant to the study is not in a repository (deidentified participant data).

## Patient and public involvement

A patient was a member of the steering committee and agreed on the study design.

## References

1. Huang C, Wang Y, Li X, et al. Clinical features of patients infected with 2019 novel coronavirus in Wuhan, China. The Lancet 2020;395:497–506 doi:10.1016/S0140-6736(20)30183-5

2. World Health Organization. Naming the coronavirus disease (COVID-19) and the virus that causes it. https://www.who.int/emergencies/diseases/novel-coronavirus-2019/technical-guidance/naming-the-coronavirus-disease-(covid-2019)-and-the-virus-that-causes-it. (accessed May 5, 2020).

3. WHO Director-General’s opening remarks at the media briefing on COVID-19 - 11 March 2020. World Health Organization Web site. https://www.who.int/dg/speeches/detail/who-director-general-s-opening-remarks-at-the-media-briefing-on-covid-19---11-march-2020.(accessed May 5, 2020).

4. COVID-19 situation update worldwide. European Centre for Disease Prevention and Control Web site. https://www.ecdc.europa.eu/en/geographical-distribution-2019-ncov-cases. (accessed May 13, 2020).

5. Myers L, Balakrishnan S, Reddy S, et al. Coronavirus Outbreak: Is Radiology Ready? Mass Casualty Incident Planning. J Am Coll Radiol Published Online First: 1 April 2020. doi:10.1016/j.jacr.2020.03.025

6. Ai T, Yang Z, Hou H, et al. Correlation of Chest CT and RT-PCR Testing in Coronavirus Disease 2019 (COVID-19) in China: A Report of 1014 Cases. Radiology Published Online First: February 26, 2020. doi:10.1148/radiol.2020200642

7. Zhao W, Zhong Z, Xie X, et al. Relation Between Chest CT Findings and Clinical Conditions of Coronavirus Disease (COVID-19) Pneumonia: A Multicenter Study. AJR Am J Roentgenol 2020;214:1072–7 doi:10.2214/AJR.20.22976

8. Li Y, Xia L. Coronavirus Disease 2019 (COVID-19): Role of Chest CT in Diagnosis and Management. AJR Am J Roentgenol Published Online First: March 4, 2020. doi:10.2214/AJR.20.22954

9. Revel M-P, Parkar A, Prosch H, et al. COVID-19 patients and the radiology department – advice from the European Society of Radiology (ESR) and the European Society of Thoracic Imaging (ESTI). Eur Radiol Published Online First: April 20, 2020. doi:10.1007/s00330-020-06865-y

10. Corman VM, Landt O, Kaiser M, et al. Detection of 2019 novel coronavirus (2019-nCoV) by real-time RT-PCR. Euro Surveill 2020;25(3) doi:10.2807/1560-7917.ES.2020.25.3.2000045

11. RC Team 2009. R: A language and environment for statistical computing. https://www.r-project.org/ R Foundation for Statistical Computing. Vienna; 2012.

12. Shi H, Han X, Jiang N, et al. Radiological findings from 81 patients with COVID-19 pneumonia in Wuhan, China: a descriptive study. Lancet Infect Dis 2020;20(4):425–434 doi:10.1016/S1473-3099(20)30086-4

13. CDC COVID-19 Response Team, Bialek S, et al. Severe Outcomes Among Patients with Coronavirus Disease 2019 (COVID-19) — United States, February 12–March 16, 2020. MMWR Morb Mortal Wkly Rep 2020;69(12):343–6 doi:10.15585/mmwr.mm6912e2

14. Chen N, Zhou M, Dong X, et al. Epidemiological and clinical characteristics of 99 cases of 2019 novel coronavirus pneumonia in Wuhan, China: a descriptive study. Lancet 2020;395(10223):507–13 doi:10.1016/S0140-6736(20)30211-7

15. Song F, Shi N, Shan F, et al. Emerging 2019 Novel Coronavirus (2019-nCoV) Pneumonia. Radiology 2020;295(1):210–7 doi:10.1148/radiol.2020200274

16. Loong T-W. Understanding sensitivity and specificity with the right side of the brain. BMJ 2003;327(7417):716–9 doi:10.1136/bmj.327.7417.716

17. Xie X, Zhong Z, Zhao W, et al. Chest CT for Typical 2019-nCoV Pneumonia: Relationship to Negative RT-PCR Testing. Radiology Published Online First: February 12, 2020. doi:10.1148/radiol.2020200343

18. Kanne JP, Little BP, Chung JH, et al. Essentials for Radiologists on COVID-19: An Update—Radiology Scientific Expert Panel. Radiology Published Online First: February 27, 2020. doi:10.1148/radiol.2020200527

19. Fang Y, Zhang H, Xie J, et al. Sensitivity of Chest CT for COVID-19: Comparison to RT-PCR. Radiology Published online February 19, 2020. doi:10.1148/radiol.2020200432

20. Tu H, Tu S, Gao S, et al. The epidemiological and clinical features of COVID-19 and lessons from this global infectious public health event. J Infect Published Online First: 18 April 2020. doi:10.1016/j.jinf.2020.04.011

21. Long C, Xu H, Shen Q, et al. Diagnosis of the Coronavirus disease (COVID-19): rRT-PCR or CT? Eur J Radiol 2020;126:108961 doi:10.1016/j.ejrad.2020.108961

22. Novel Coronavirus Pneumonia Emergency Response Epidemiology Team. The epidemiological characteristics of an outbreak of 2019 novel coronavirus diseases (COVID-19) in China. Zhonghua Liu Xing Bing Xue Za Zhi 2020;41:145–51 doi:10.3760/cma.j.issn.0254-6450.2020.02.003

23. Chen G, Wu D, Guo W, et al. Clinical and immunological features of severe and moderate coronavirus disease 2019. J Clin Invest 2020;130:2620–9 doi:10.1172/JCI137244

24. Liu K-C, Xu P, Lv W-F, et al. CT manifestations of coronavirus disease-2019: A retrospective analysis of 73 cases by disease severity. Eur J Radiol 2020;126:108941 doi:10.1016/j.ejrad.2020.108941

25. Bernheim A, Mei X, Huang M, et al. Chest CT Findings in Coronavirus Disease-19 (COVID-19): Relationship to Duration of Infection. Radiology Published Online First: February 20, 2020. doi:10.1148/radiol.2020200463

26. Lovato A, de Filippis C. Clinical Presentation of COVID-19: A Systematic Review Focusing on Upper Airway Symptoms. Ear Nose Throat J Published Online First: April 13, 2020. doi:10.1177/0145561320920762

27. Zhang J-J, Dong X, Cao Y-Y, et al. Clinical characteristics of 140 patients infected with SARS-CoV-2 in Wuhan, China. Allergy Published Online First: 19 February 2020. doi:10.1111/all.14238

